# COVID-19 induced birth sex ratio changes in England and Wales

**DOI:** 10.1101/2022.09.09.22279763

**Authors:** Gwinyai Masukume, Margaret Ryan, Rumbidzai Masukume, Dorota Zammit, Victor Grech, Witness Mapanga, Yosuke Inoue

## Abstract

**Background:** The sex ratio at birth (male live births divided by total live births) may be a sentinel health indicator. Stressful events reduce this ratio 3-5 months later by increasing male fetal loss. This ratio can also change 9 months after major population events that are linked to an increase or decrease in the frequency of sexual intercourse at the population level, with the ratio either rising or falling respectively after the event. We postulated that stress caused by the COVID-19 pandemic may have affected the ratio in England and Wales.

**Methods:** Publicly available, monthly live birth data for England and Wales was obtained from the Office for National Statistics up to December 2020. The sex ratio at birth for 2020 (global COVID-19 onset) was predicted using data from 2012-2019. Observed and predicted values were compared.

**Results:** Three months after COVID-19 was declared pandemic (March 2020), there was a significant fall in the sex ratio at birth to 0.5100 in June 2020 which was below the 95% prediction interval of 0.5102-0.5179. Nine months after the pandemic declaration, (December 2020), there was a significant rise to 0.5171 (95% prediction interval 0.5085-0.5162). However, December 2020 had the lowest number of live births of any month from 2012 to 2020.

**Conclusions:** Given that June 2020 falls within the crucial window when population stressors are known to affect the sex ratio at birth, these findings imply that the start of the COVID-19 pandemic caused population stress with notable effects on those who were already pregnant by causing a disproportionate loss of male fetuses. The finding of a higher sex ratio at birth in December 2020, i.e., 9 months after COVID-19 was declared a pandemic, suggests that lockdown restrictions initially spurred more sexual activity in a subset of the population in March 2020.

## Introduction

Male live births outnumber female live births at the population level (Graunt 1676). The secondary sex ratio, commonly known as the sex ratio at birth (SRB), is calculated as male divided by total live births (Grech 2014). The SRB may serve as a sentinel health indicator, revealing unfavourable conditions through a decline and better conditions through an increase (Davis et al. 1998; Grech & Masukume 2016).

In response to population-level events, the SRB may remain unchanged (Grech & Scherb 2021; Masukume & Grech 2016), transiently increase, or decrease. The increases or decreases have been noted to occur within two distinct time windows. The first window is 3-5 months after sudden and unanticipated stressful events such as terrorist attacks (Bruckner et al. 2010), the death of a well-known public person (Grech 2015), or unexpected national election results (Retnakaran & Ye 2020). This first window is linked to a disproportionate pregnancy loss of male fetuses, which is reflected in a lower SRB a few months after the stressful event. Male fetuses are more vulnerable to the effects of maternal stress (Aiken & Ozanne 2013; Catalano et al. 2021), in accordance with the Trivers-Willard hypothesis, which holds that unfavourable environmental conditions might lower the ratio of males to females (Trivers & Willard 1973).

The second window occurs 9 months after an event and can result in either a drop or a rise in the SRB. The SRB can decrease 9 months after a stressful event, such as a massive earthquake, and this is ascribed to a decrease in population-level sexual intercourse frequency and/or reduced sperm motility (Fukuda et al. 2018; Fukuda et al. 1996). The SRB may increase 9 months later if the event is linked to a population-level opportunity for more frequent sexual activity, such as the celebratory atmosphere during the first-ever home Fédération Internationale de Football Association (FIFA) 2010 World Cup tournament with a strong showing by the local team (Masukume & Grech 2015). It has been established that SRB and the fertile phase of the menstrual cycle have a U-shaped association (Guerrero 1974). The SRB would be skewed toward male births because of this U-shaped association if coitus was more common at the population level and more conceptions occurred further from the centre of the fertile period.

It has been suggested that an SRB decrease could be brought on by COVID-19 stress (Abdoli 2020). Indeed, we noted that the SRB fell in South Africa in June 2020, 3 months after COVID-19 was proclaimed a pandemic (Masukume et al. 2022), and in Japan in December 2020, 9 months after the proclamation (Inoue & Mizoue 2022).

March 2020 was a key month for COVID-19 in England and Wales. This month saw an increase in local and international media coverage of the virus (Ng et al. 2021) as the World Health Organization (WHO) declared COVID-19 to be a pandemic (Cucinotta & Vanelli 2020). The first COVID-19 related death in England was also reported in March 2020 (Mahase 2020). Furthermore, on 23 March 2020, the Prime Minister of the United Kingdom (UK), i.e., England, Wales, Scotland and Northern Ireland made a major announcement on ‘lockdown’ restrictions, the closure of non-essential enterprises and the requirement to stay at home save for a few limited circumstances, in one of the most watched broadcasts. Higher levels of COVID-19-related anxiety, depression and trauma symptoms were observed in the general population in the UK from 23 to 28 March 2020 compared to previous epidemiologic periods (Shevlin et al. 2020). This study sought to ascertain whether the SRB in England and Wales changed in response to the COVID-19 pandemic, as well as when this change took place.

## Material and methods

### Data and statistical analysis

We used publicly available, monthly live birth data for England and Wales obtained from the Office for National Statistics, which is typically the most comprehensive data source available (Office for National Statistics 2022). The 108-month period (9 years) covered by the data, from January 2012 to December 2020, was consistent with the number of months examined in earlier research on the subject (Inoue & Mizoue 2022; Masukume et al. 2022). The most recent month for which data was available was December 2020.

The source of live birth data is a legal record that is the result of a child’s parent(s) or informant formally registering the birth at a registry office. The doctor or midwife who attended the birth fills out a birth notification form. The coronavirus pandemic did not influence birth notifications the same way it did on birth registrations, which were delayed. For 2020, 0.3% of birth registrations across England and Wales were still unlinked to birth notifications (Office for National Statistics 2021).

Using data from January 2012 to December 2019, we predicted SRBs for the 2020 months because the disease now known as COVID-19 was initially notified to WHO on 31 December 2019 (Wu & McGoogan 2020). We utilized an autoregressive moving average (ARMA) model with the minimum Akaike Information Criterion (AIC) (i.e., autoregressive parameter [4] and moving average parameter [4]) to fit SRBs for January 2012 to December 2019 after checking for stationarity with the Dickey-Fuller test (p < .001) (Table S1). The results from this model were applied to predict SRBs for 2020. The 95% upper and lower bounds of the predicted SRBs were calculated using Stata’s ‘predict’ command and the ‘mse’ option (StataCorp 2019).

### Ethical considerations

The data were anonymized, thus ethical approval was not required.

## Results

A total of 6,108,030 live births—3,133,915 males and 2,974,115 females—were recorded during the 9-year period between January 2012 and December 2020, with an SRB of 0.5131. From 2012 through 2020, December 2020 had the fewest live births of any month with 47,291 (Figure 1). The SRB for June 2020 was 0.5100, the lowest of any June throughout the study period. June 2020 was also the first time June had had the lowest SRB of the year. The SRB for December 2020 was 0.5171, which was the highest for any December throughout the study period. Additionally, December 2020 marked the first time the month had the highest SRB of the year (Figure 2). The lowest yearly SRB over the 9-year period was 0.5122 observed in 2020.

**Figure 1.**
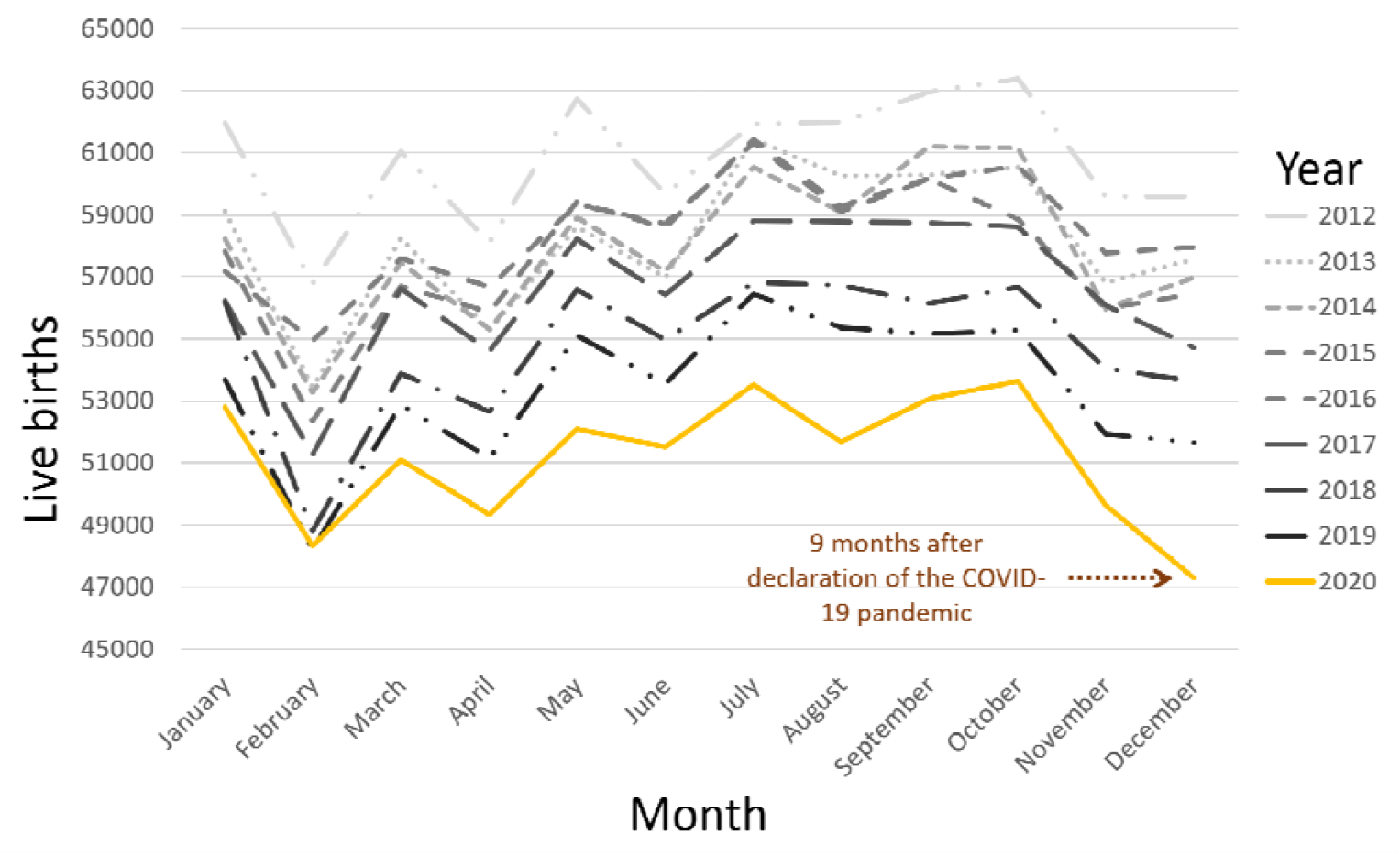
Total live births over 9 years, from January 2012 to December 2020.

**Figure 2.**
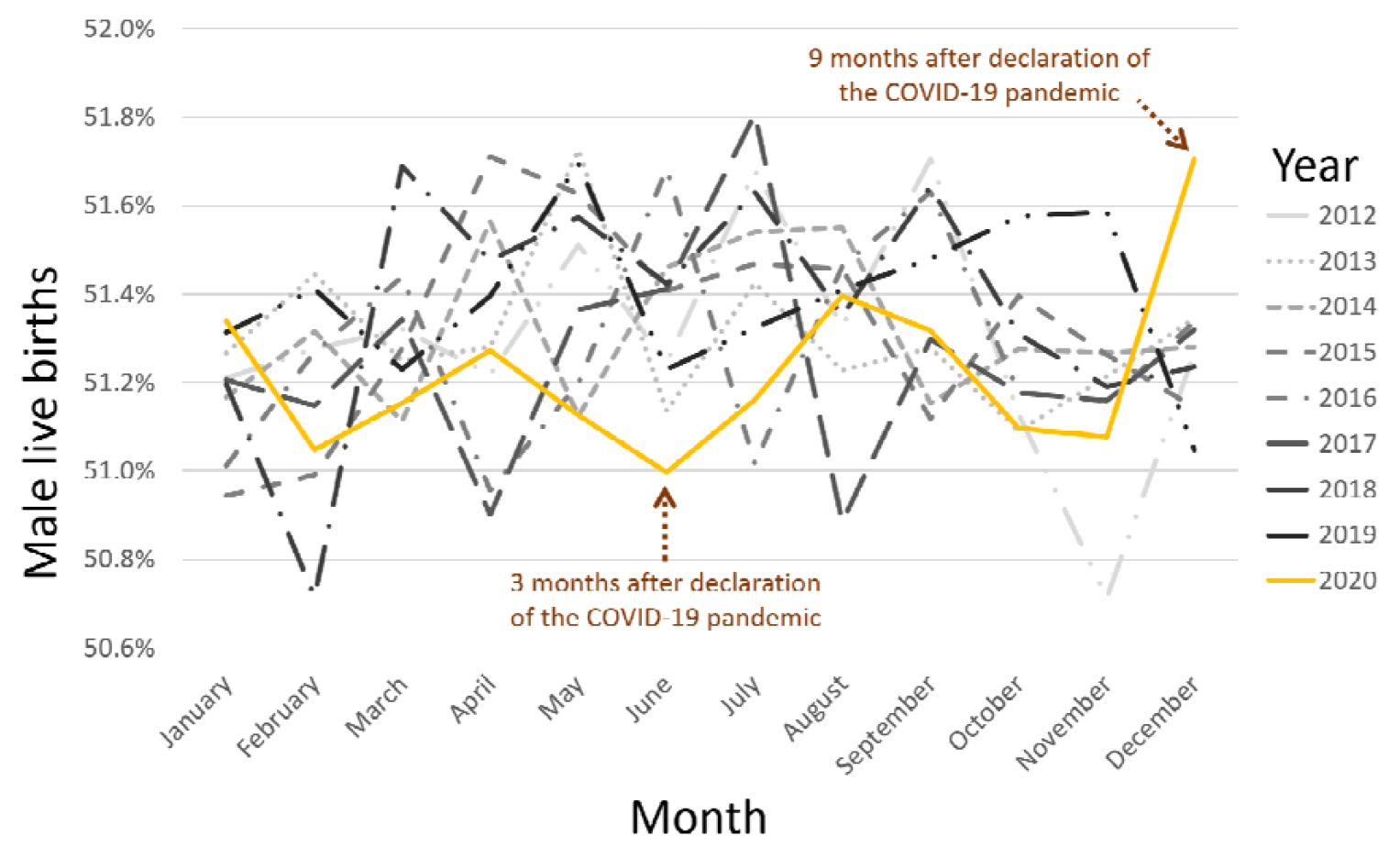
Monthly proportion of male live births over 9 years, from January 2012 to December 2020.

There was a significant fall in the SRB of 0.5100 in June 2020 which was below the 95% prediction interval of 0.5102-0.5179. In December 2020, there was a significant rise in the SRB of 0.5171 which was above the 95% prediction interval of 0.5085-0.5162 (Figure 3 and Table 1).

**Table 1.**
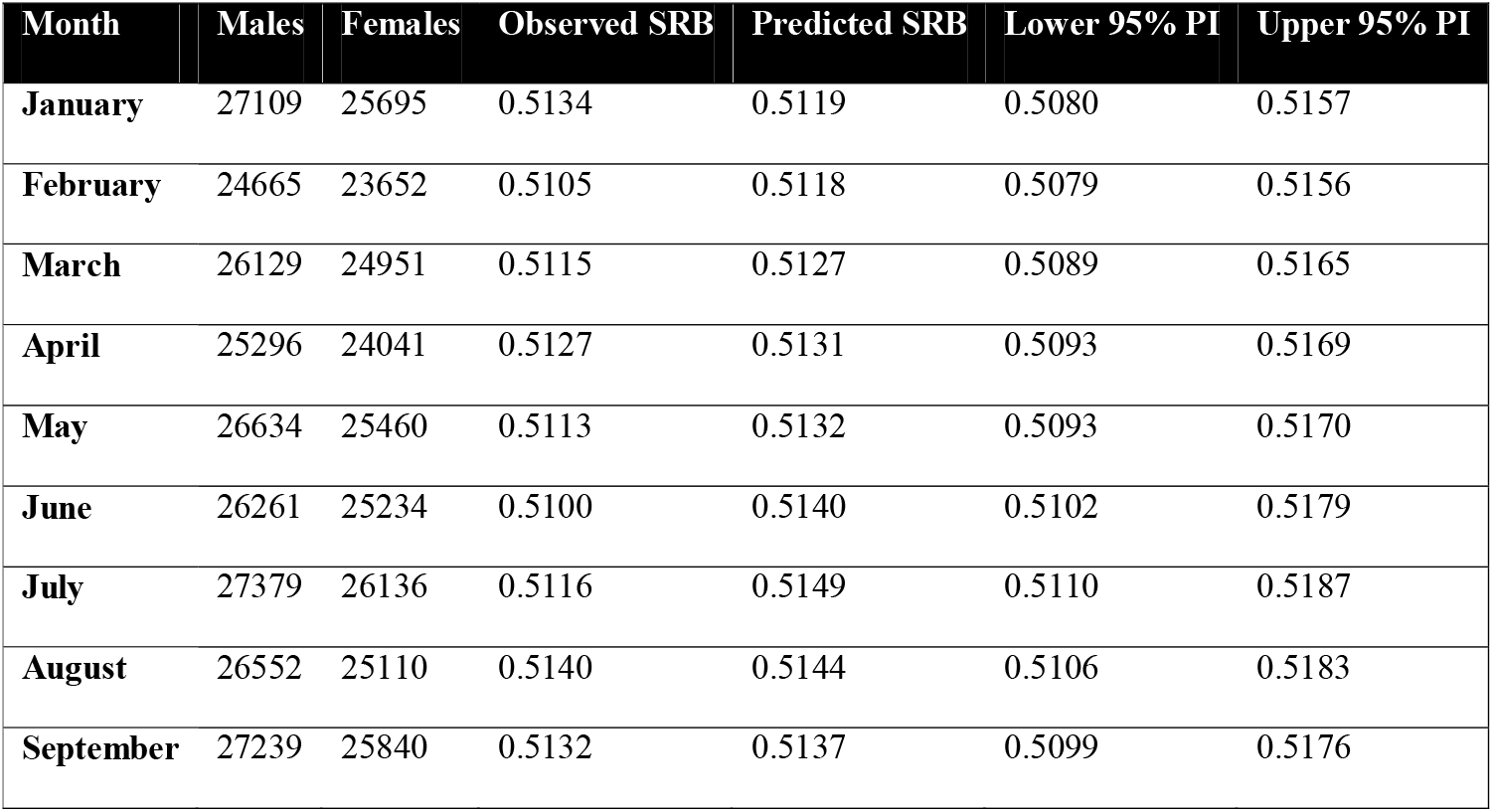
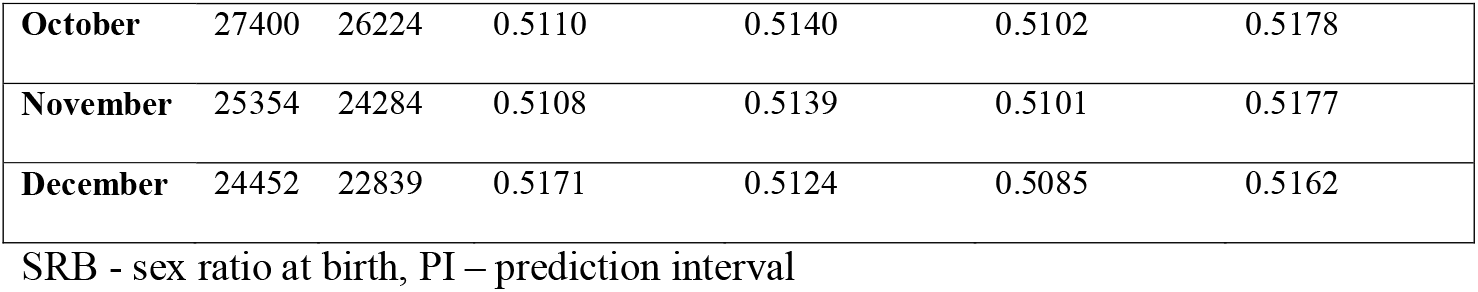
Observed and predicted sex ratio at birth for 2020.

**Figure 3.**
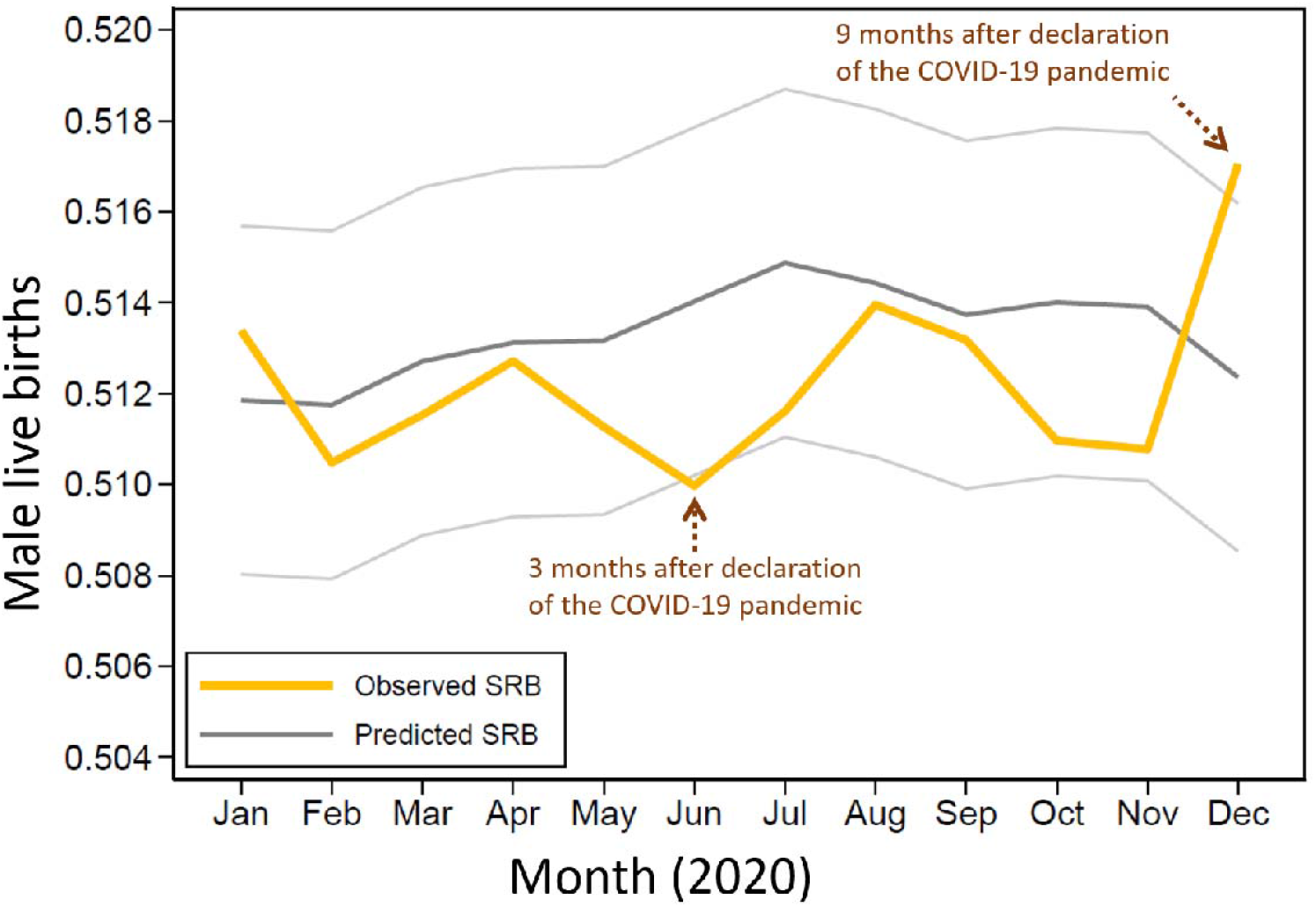
Observed and predicted sex ratio at birth (SRB) in 2020.

## Discussion

### Principal findings

In this study, we observed that the COVID-19 pandemic affected SRB in England and Wales. We observed that December 2020 had notably the fewest live births of any month over the 108-month study period, with 47,291 live births, 9 months after the pandemic was declared in March 2020. With an SRB of 0.5100, June 2020’s SRB was significantly the lowest of any June during the study period. Indeed, the lowest SRB of the year was recorded for the first time in June 2020. This SRB decline took place 3 months after the pandemic was declared. The December 2020 SRB, which was 0.5171, was significantly the highest SRB for any December throughout the study period. Indeed, for the first time in 2020, December saw the highest SRB of the year.

### Comparison with other studies

Three months after the COVID-19 pandemic was declared, a decline in the SRB was noted in South Africa in June 2022 (Masukume et al. 2022), which is similar to the transient decline in SRB observed in this study in June 2020. This is consistent with studies that found a transient reduction in SRB 3-5 months after unanticipated occurrences that stressed populations (Calleja 2020; Catalano et al. 2006; Grech 2015; Retnakaran & Ye 2020).

Excess male fetal loss, particularly in those who were 20 to 28 weeks (4-7 months) pregnant at the time of the major stressful event, has been linked to a drop in the SRB 3 months later (Bruckner et al. 2010). A stillbirth is defined in England and Wales as a fetal death that occurs at ≥ 24 completed weeks of pregnancy, whereas a miscarriage happens at < 24 completed weeks (Gurol-Urganci et al. 2022; Jones et al. 2022). Thus, our findings indicating a decreased SRB in England and Wales in June 2020, 3 months after the declaration of the COVID-19 pandemic, imply an increase in miscarriage and stillbirth rates during the early part of the COVID-19 pandemic, disproportionately affecting male fetuses.

Moreover, a rise in stillbirths from 2.38 per 1,000 births between 1 October 2019 and 31 January 2020 (defined as the pre-pandemic period) to 9.31 per 1,000 births between 1 February and 14 June 2020 (defined as the pandemic period) was reported from an English centre (Khalil et al. 2020). Although the sex of the stillbirths was not provided, in this English paper, male fetuses typically account for the majority of stillbirths (Mondal et al. 2014). These particular stillbirths had no connection to clinically severe acute respiratory syndrome coronavirus 2 (SARS-CoV-2) infection, which indicates an indirect stillbirth mechanism possibly involving increased maternal stress and anxiety. Internationally, miscarriages are typically not included in such statistics (Gurol-Urganci et al. 2022; Jones et al. 2022). Because a drop in the SRB might also be used to detect male fetal loss (Bruckner et al. 2010), our results also indicate that there may have been an unrecognised excess of male fetal loss before 24 weeks gestation (miscarriages). This is because national level statistics from England (Gurol-Urganci et al. 2022) and Wales (Jones et al. 2022) did not indicate a rise in stillbirths throughout the COVID-19 period of the current investigation, in contrast to the aforementioned single centre study (Khalil et al. 2020).

Despite the SRB rising, in England and Wales, in December 2020, 9 months after the COVID-19 pandemic was declared, this happened in the context of a sharp reduction in overall live births compared to Decembers prior. Contrasting this with the fact that 9 months following the 2010 FIFA World Cup in South Africa, there was an increase in SRB along with an increase in the overall number of live births (Masukume & Grech 2015; Masukume et al. 2016). This suggests that whereas a greater fraction of the population engaged in sexual activity more frequently in South Africa during the 2010 World Cup, a smaller portion of the population did so in England and Wales in March 2020. This is consistent with a British study that looked at the first 4 months of the lockdown and found that some people who were cohabiting felt like they had more partnered sexual activity since the lockdown, but people who were not cohabiting felt like they had less (Mercer et al. 2021).

Contrary to our current findings, the SRB fell in Japan, as we previously reported, in December 2020, 9 months after COVID-19 was designated a pandemic (Inoue & Mizoue 2022), but this Japanese decline might have been within the same context as a significant decline in overall live births for December 2020 seen in England and Wales (Ghaznavi et al. 2022). This would suggest less frequent sexual encounters (Guerrero 1974), and indeed declines in sexual activity associated with the start of the COVID-19 pandemic have been suggested in Japan (Kitamura et al. 2021).

Overall, our research demonstrates that the SRB is a very valuable sentinel health indicator since, unlike other indicators, it is usually available for entire populations (Davis et al. 1998). Population dynamics can be better understood by triangulating the SRB with additional data.

### Strengths and limitations

The study’s results cannot be extrapolated to the individual level due to the ecological fallacy (Björk et al. 2021). However, the ecologic study method is arguably the most appropriate for examining how a population stressor affects a population outcome (Pearce 2011).

March 2020 saw the temporary suspension of birth registration in England and Wales. As soon as it was safe to do so, registrations started up again in June 2020. Under normal circumstances, more than 95% of births are typically documented within the required 42-day window. In 2020, only 58% of births were reported within 42 days, a notable reduction. Due to this known delay, the cut-off date for incorporating 2020 live birth registrations was five and a half months later than usual, in August 2021. Similar distributions for the baby’s sex were found in both the linked registrations data and the unlinked notifications when birth registrations and birth notifications were compared. The aforementioned birth registration delays had little bearing on the current analysis because 0.3% of notifications across England and Wales were still unlinked (Office for National Statistics 2021).

The analysis presented here pertains to England and Wales’ initial COVID-19 wave (Knock et al. 2021). England and Wales had seen more waves at the time of writing. In the waves that followed, the SRB might have been perturbed. When the live birth data for these times are made accessible, this is a topic for further study. In the years between 2012 and 2020, we also noticed other major SRB dips and peaks, but we had not *a priori* hypothesised how they might have arisen; thus, we did not address them further in this paper.

### Conclusions

These results suggest that the start of the COVID-19 pandemic caused population stress with notable effects on those who were already pregnant by causing a disproportionate loss of male fetuses given that June 2020 falls within the critical window when population stressors are known to affect the sex ratio at birth. The finding of a higher sex ratio at birth in December 2020, 9 months after COVID-19 was declared a pandemic, indicates that lockdown regulations initially encouraged more partnered sexual activity in a small portion of the population. Future pandemic preparedness (Marston et al. 2017) and social policy (Cook & Ulriksen 2021) can be influenced by our findings. Examples of how this knowledge could be applied to prevent a potential SRB reduction include targeted increases in resources for maternal health services, including improved fetal surveillance, particularly during and in the lead up to 3-5 months after the start of a future pandemic. Future studies should examine whether the SRB changed in other nations after the onset of COVID-19.

## Data Availability

All data produced are available online at https://www.ons.gov.uk/peoplepopulationandcommunity/birthsdeathsandmarriages/livebirths/adhocs/15002livebirthsbymonthofoccurrenceandsexofbabyenglandandwales2012to2020

## Acknowledgements

We acknowledge the Office for National Statistics for the recorded live birth data.

## Author Contributions

Conceptualization: GM, MR, RM, DZ, VG, WM, YI Data curation: VG, GM, YI

Formal analysis: YI, VG, GM Writing – original draft: GM

Writing – review & editing: MR, RM, GM, DZ, VG, WM, YI

---

**Table S1.** Parameter selection procedure for the autoregressive moving average (ARMA) model to fit and predict secondary sex ratio in England d Wales.

| AR | MA | AR |  |  |  |  | MA |  |  |  |  | AIC |
| --- | --- | --- | --- | --- | --- | --- | --- | --- | --- | --- | --- | --- |
|  |  | L1 | L2 | L3 | L4 | L5 | L1 | L2 | L3 | L4 | L5 |  |
| 1 | 0 | -0.01 |  |  |  |  |  |  |  |  |  | -897.15 |
| 1 | 1 | 0.92 |  |  |  |  | -1.00 |  |  |  |  | -897.39 |
| 1 | 2 | 0.91 |  |  |  |  | -0.96 | -0.04 |  |  |  | -897.52 |
| 1 | 3 | 0.89 |  |  |  |  | -0.95 | 0.01 | -0.07 |  |  | -893.89 |
| 1 | 4 | 0.71 |  |  |  |  | -0.75 | 0.02 | -0.03 | -0.07 |  | -891.4 |
| 1 | 5 | 0.62 |  |  |  |  | -0.67 | 0.01 | 0.04 | 0.05 | -0.21 | -891.78 |
| 2 | 0 | -0.01 | 0.02 |  |  |  |  |  |  |  |  | -895.18 |
| 2 | 1 | 0.95 | -0.04 |  |  |  | -1.00 |  |  |  |  | -897.54 |
| 2 | 2 | -0.01 | 0.86 |  |  |  | -0.10 | -0.90 |  |  |  | -896.00 |
| 2 | 3 | NC |  |  |  |  | NC |  |  |  |  |  |
| 2 | 4 | NC |  |  |  |  | NC |  |  |  |  |  |
| 2 | 5 | 1.30 | -0.77 |  |  |  | -1.44 | 0.95 | -0.01 | -0.01 | -0.17 | -899.75 |
| 3 | 0 | -0.01 | 0.02 | -0.02 |  |  |  |  |  |  |  | -893.2 |
| 3 | 1 | 0.95 | 0.02 | -0.07 |  |  | -1.00 |  |  |  |  | -896.00 |
| 3 | 2 | -1.53 | -0.99 | -0.05 |  |  | 1.57 | 1.00 |  |  |  | -893.94 |
| 3 | 3 | NC |  |  |  |  | NC |  |  |  |  |  |
| 3 | 4 | -0.08 | -0.02 | -0.74 |  |  | 0.02 | 0.09 | 0.88 | 0.16 |  | -894.88 |
| 3 | 5 | 2.23 | -2.03 | 0.75 |  |  | -2.40 | 2.41 | -0.94 | -0.18 | 0.12 | -898.86 |
| 4 | 0 | -0.01 | 0.02 | -0.02 | -0.01 |  |  |  |  |  |  | -891.21 |
| 4 | 1 | 0.72 | 0.02 | -0.03 | -0.11 |  | -0.75 |  |  |  |  | -892.38 |
| 4 | 2 | -0.82 | -0.92 | 0.06 | 0.08 |  | 0.83 | 0.97 |  |  |  | -891.28 |
| 4 | 3 | 1.90 | -1.71 | 0.68 | -0.13 |  | -2.65 | 2.75 | -1.35 |  |  | -899.42 |
| 4 | 4 | 1.71 | -1.83 | 1.51 | -0.88 |  | -1.80 | 2.05 | -1.80 | 1.00 |  | -904.70 |
| 4 | 5 | 1.39 | 0.05 | -1.20 | 0.65 |  | -1.23 | 0.03 | 1.05 | -0.68 | -0.16 | -897.78 |
| 5 | 0 | -0.01 | 0.01 | -0.01 | -0.02 | -0.18 |  |  |  |  |  | -892.38 |
| 5 | 1 | 0.56 | 0.02 | -0.02 | -0.01 | -0.2 | -0.61 |  |  |  |  | -893.52 |
| 5 | 2 | NC |  |  |  |  | NC |  |  |  |  |  |
| 5 | 3 | NC |  |  |  |  | NC |  |  |  |  |  |
| 5 | 4 | 0.73 | -0.37 | 0.79 | -0.82 | -0.18 | -0.90 | 0.47 | -0.84 | 0.96 |  | -903.07 |
| 5 | 5 | NC |  |  |  |  | NC |  |  |  |  |  |

AIC: Akaike Information Criterion; AR: autoregressive parameter; MA: moving average parameters; NC: not converged.

